# Beyond Citations: Identifying Transformational Research in Hypospadias Through Bibliometrics and the Disruption Index

**DOI:** 10.64898/2026.04.18.26351160

**Authors:** Tariq Abbas, Mansura Naznine, Mikha Mykha, Maraeh Mancha, Anushka Hardas, Putu Angga Risky Raharja, Muhammad E. H. Chowdhury

## Abstract

Hypospadias, a common congenital anomaly requiring surgical correction, has seen growing research in surgical techniques and outcomes. However, no comprehensive bibliometric or disruption-based analysis exists to map the field’s evolution. This study uses bibliometrics and the Disruption Index (DI) to identify key transformational research in hypospadias. A systematic search of five databases (PubMed, Web of Science, ScienceDirect, Scopus, and Dimensions) from January 1990 to December 2023 was conducted, yielding 7,732 articles. After applying inclusion criteria, 200 studies were analyzed. Citation data and DI scores were calculated using OpenCitations. Spearman’s rank test assessed correlations between DI and citation metrics. A subgroup analysis identified trends based on the latest hypospadias research priorities. The mean citation count was 72.3 (SD = 43.1) with a mean DI of 0.011 (SD = 0.17). Five studies, focusing on complications, analgesia, and surgical techniques, had the highest DI (1.0). A moderate positive correlation was found between DI and citation rate (ρ = 0.405, p < 0.001). Subgroup analysis showed most research focused on surgical techniques (30.5%) and etiology (25.8%), while areas like surgical training (2.6%) and innovation (0%) were underrepresented. This study identifies critical gaps in hypospadias research. The DI reveals influential studies that redirect research trajectories. Future work should focus on innovation and translational research to accelerate advancements in hypospadias care.

## 1. INTRODUCTION

Hypospadias is a common congenital anomaly of the male external genitalia characterized not only by an abnormally positioned urethral meatus along the ventral aspect of the penis, but also by varying degrees of ventral penile curvature (chordee), dorsal hooded prepuce, and underlying hypoplasia of the penile shaft tissues. This complex phenotype has implications for both functional and cosmetic surgical outcomes, [Abbas & McCarthy, 2016] often requires surgical correction to ensure normal urinary function and, in many cases, to achieve aesthetic and sexual function in adulthood.[Spinoit et al, 2021] Urethroplasty, the surgical procedure employed to correct hypospadias, has evolved significantly over the past few decades, with a variety of surgical techniques introduced to address the complex nature of the malformation. However, despite these advancements, the management of hypospadias remains a challenging and highly specialized field within pediatric urology, with significant variability in surgical outcomes and long-term follow-up results.[Abbas et al, 2023]

Over the last two decades, a growing body of literature has emerged on the topic of hypospadias surgery, focusing on novel surgical techniques, innovations in surgical tools and approaches, genetic etiologies, artificial intelligence, tissue engineering, patient outcomes, and the training of urologic surgeons. [Abbas 2017, 2025; Khondkher et al, 2025] These studies have not only contributed to the refinement of surgical techniques but have also emphasized the importance of individualized treatment plans and improving the quality of life for patients. Furthermore, various studies have highlighted the need for ongoing professional education and training to ensure that surgeons are equipped with the most up-to-date knowledge and skills in performing hypospadias surgery.[Bhatia et al, 2024]

Despite the abundance of research, a comprehensive Bibliometric analysis, which evaluate the impact and trends of research through citation and metadata analysis is lacking and could provide valuable insights into the evolving landscape of Hypospadiology field. These analyses can highlight shifts in research focus, the influence of specific studies, and emerging areas of interest that require further investigation. A bibliometric approach also allows for the identification of influential studies, authors, and journals, thereby offering a deeper understanding of how research in hypospadias and urethroplasty has developed over time. [Manoj Kumar, George, and Anisha, 2023] In addition to traditional bibliometric approaches such as citation counts and co-authorship networks, recent advances have introduced the concept of disruption-based analysis. The Disruption Index (DI), first proposed by Wu et al., measures the extent to which a publication disrupts or consolidates existing knowledge. A highly disruptive article is one that leads future research to ignore the prior work it builds upon, thereby shifting the direction of the field. In contrast, consolidating papers amplify existing paradigms. These metric complements citation analysis by offering insights into the transformative potential of scientific contributions, rather than their popularity or reach alone.

This study conducts a bibliometric analysis of hypospadias and urethroplasty research from 2000 to 2023, assessing trends in surgical techniques, outcomes, and training innovations. We aim to evaluate the academic impact of key studies, identify knowledge gaps, and propose future research directions. We hypothesize that combining traditional citation metrics with the Disruption Index (DI) will provide a clearer understanding of research evolution and emerging trends.

## 2. METHODOLOGY

### 2.1 Evidence Acquisition

This study adheres to the Preferred Reporting Items for Systematic Reviews and Meta-Analyses (PRISMA) guidelines, [Page et al, 2020] which provide a robust framework for systematically assessing and synthesizing research in a transparent and reproducible manner. A comprehensive literature search was conducted using five widely recognized electronic databases: PubMed, Web of Science, ScienceDirect, Scopus, and Dimensions. These databases were selected for their extensive coverage of medical, surgical, and basic science studies related to hypospadias. The search employed a Boolean strategy with the keywords: (“hypospadias”) AND (“surgery” OR “outcome” OR “training”). These terms were consistently applied across all five databases to ensure uniformity in the search process. The search was restricted to publications between January 1, 1990, and December 31, 2023, to capture recent advancements while also analyzing long-term trends in research related to hypospadias and urethroplasty.

### 2.2 Study Selection and Screening

This bibliometric study included publications on hypospadias without subtype restrictions, focusing on surgical techniques, clinical outcomes, and innovations. Systematic reviews, meta-analyses, and consensus guidelines were also included. Only English-language studies with complete metadata and accessible full texts were considered, while studies on unrelated urological conditions, conference abstracts, editorials, and duplicates were excluded. Two independent investigators screened titles, abstracts, and full texts for relevance, resolving disagreements through discussion. The selection process was documented using a PRISMA flow diagram. A total of 7,732 publications were initially added into the pre-included study to underwent further examination from these, the most relevant 2923 papers were selected for further analysis. Following the collection of the dataset, citation counts for all 1,759 papers were extracted using Scopus. Scopus was selected due to its reputation as a trustworthy citation database that provides precise and current citation metrics. The top 200 most-cited papers were determined by ranking the papers according to citation counts. [Vaidyanathan,2024] For each of the top 200 papers, the Digital Object Identifier (DOI) was extracted to ensure precise tracking of each publication. Keywords were extracted using MESH terms from Pubmed rather than relying on author-supplied keywords.

The Disruption Index could not be calculated directly using Scopus due to limited access to citing and referenced papers. To address this, OpenCitations was used to calculate the index for the top 200 most-cited papers. Citation counts from both Scopus and OpenCitations were reported to ensure transparency. All citation data were recorded on March 22, 2025. The Disruption Index evaluates the impact of an article by reviewing its citation and reference patterns, providing insights into how it influences future research [Funk and Owen-Smith,2016]. The approach began by extracting citing papers (studies mentioning the focal publication) and reference papers cited by the focal article. The Disruption Index was calculated by categorizing citing papers into three groups: 1) **disruptive citing papers** (citing the focal paper but not its references), 2) **non-disruptive citing papers** (citing both the focal paper and its references), and 3) **reference-only citing papers** (citing only the references, not the focal paper itself). [Leydesdorff and Bornmann, 2021].

The Disruption Index was computed using the following formula:

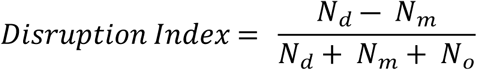

Where, *N*_*d*_ represents the number of disruptive citing papers, *N*_*m*_ represents the number of non-disruptive citing papers, and *N*_*o*_represents the number of papers that cite only the references of the focal paper. This formula quantifies how a paper disrupts research by influencing future studies. A Python-based pipeline was developed to automate data extraction from OpenCitations, compute the Disruption Index, and store results in an Excel file. Time delays were added to manage OpenCitations’ rate limits.

### 2.3 Data Extraction

Key metadata and bibliometric parameters were collected for each study, including title, authors, journal, year, and study type. A subgroup analysis assessed the proportion of research themes in hypospadiology based on the recent global Delphi survey to understand the distribution of efforts in high-impact publications.[Abbas et al, 2024]

### 2.4 Quality Assessment

The quality of the top 20 publications with the highest Disruption Index (DI) scores was assessed by two reviewers, with a third consulted to resolve discrepancies. Randomized Controlled Trials (RCTs) were evaluated using the RoB 2 tool, observational studies with the Newcastle-Ottawa Scale (NOS), and review articles with the AMSTAR 2 tool. Case reports were assessed using the JBI Checklist. Studies were categorized as good, fair, or poor based on methodological rigor, completeness, and bias. For non-standard study types, inclusion decisions were made through reviewer consensus.

### 2.5 Data Analysis

To assess the relationship between the Citation Disruption (CD) Index and traditional citation metrics (average and total citations from Scopus), correlation analyses were conducted using SPSS Version 29. The CD Index quantifies how much a publication disrupts research by influencing subsequent studies in a novel way, distinguishing between disruptive and non-disruptive citations. Spearman’s rank correlation coefficient (ρ) was used due to non-normal data distribution, with statistical significance set at p < 0.05. This analysis aimed to better characterize the transformative impact of hypospadias research.

## 3. RESULTS

A total of 200 articles were included, spanning from January 1, 1990 to December 31, 2023. The annual number of publications showed a general upward trend, with notable peaks in 2020. The total number of citations across all included articles was 18046 with a mean of 72.3 citations per article and a median of 57. The average number of citations per year per article was 4.11. These studies were selected from a pool of publications retrieved through a systematic search of five databases: PubMed, Web of Science, ScienceDirect, Scopus, and Dimensions. **(Figure 1)**

**Figure 1.**
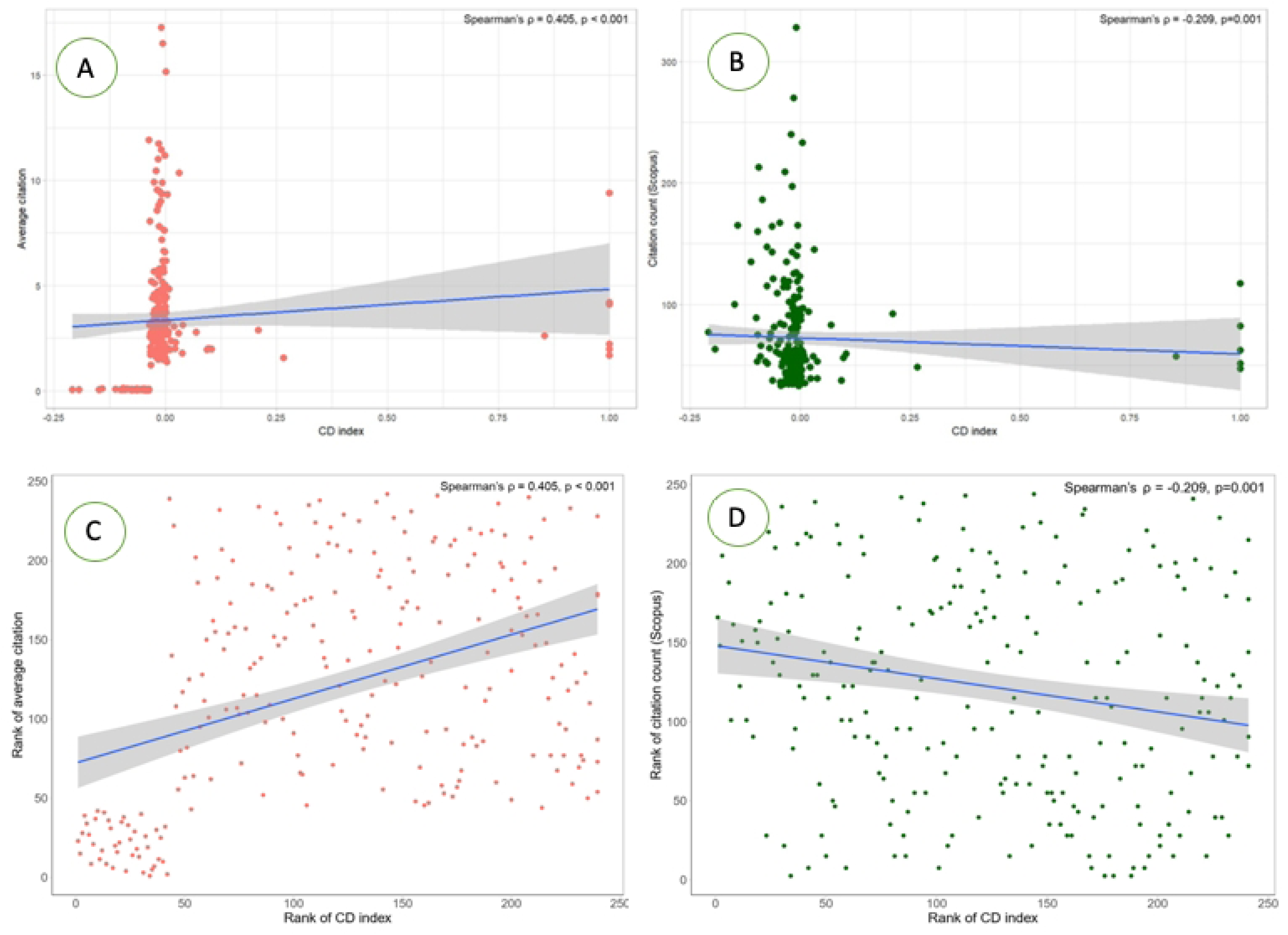
PRISMA Flow Diagram.

### 3.1 Bibliometric Overview

The mean citation count across the entries was 72.3, with a standard deviation of 43.1, indicating a moderate level of variability in the total number of citations. The mean average citations per article was 7.3, with a standard deviation of 2.45, suggesting that individual articles tended to receive citations in the range of 5 to 10, though some articles were cited more frequently. The mean disruption index (CD) was 0.011, with a standard deviation of 0.17. Bibliometric details of the Top 20 publications with the highest DI is summarized in Table 1.

### 3.2 Correlation between the CD index and Scopus citation count

Correlation between CD Index and Average citation (average number of citations per year since publication) and Citation Count from Scopus were conducted resulting in a moderate positive correlation between the CD index and average citation count (ρ = 0.405, p < 0.001), indicating that a higher CD index is associated with a higher average citation (total number of citations per number of publication). (**Figure 2, A**) And, there is a weakly significant negative correlation between the CD index and Scopus citation count (ρ = −0.209, p = 0.001), indicating that a higher CD index is associated with a lower Scopus citation count. (**Figure 2, B**)

**Figure 2.**
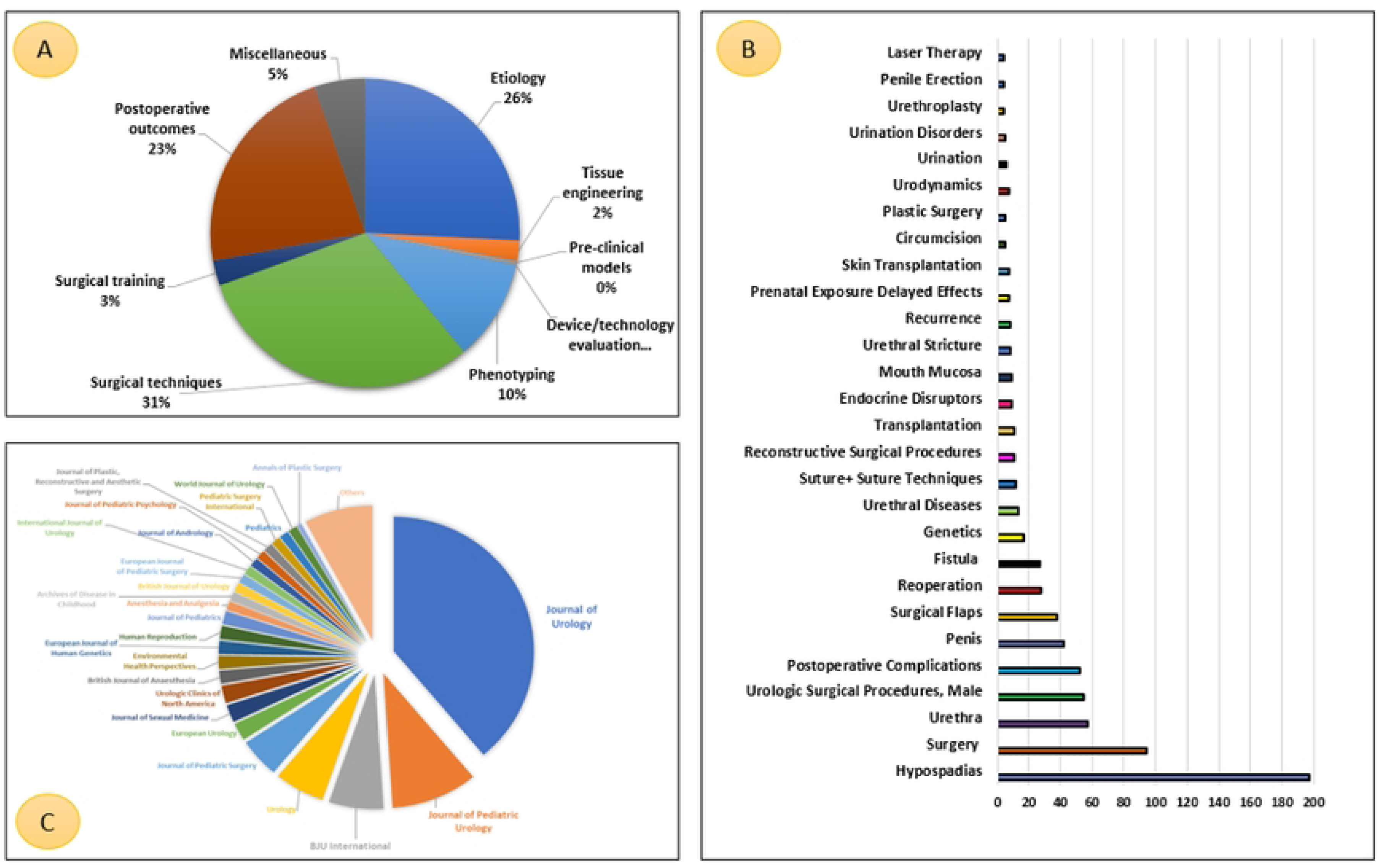
Correlation between the CD Index and Average Citation (A) and Citation Count (Scopus) (B). (C) Ranked Correlation analysis between Average citation and CD Index. (D) Ranked Correlation analysis between Citation count (Scopus) and CD Index.

There is a statistically significant moderate positive correlation (Spearman’s ρ = 0.405, p < 0.001) between the rank of the CD index and the rank of average citation count. Higher CD index ranks (higher CD index) are associated with higher average citation count ranks (higher average citation). (**Figure 2, C**) On the other hand, there is a statistically significant but weak negative correlation (Spearman’s ρ = –0.209, p = 0.001) between the rank of the CD index and the rank of citation count in Scopus. Higher CD index ranks (higher CD index) are associated with lower ranks of Scopus citation count ranks (fewer citations). The scattered distribution of the dots suggests the weak correlation. (**Figure 2, D**)

### 3.3 Subgroup Analyses

Subgroup analyses were conducted based on the study type, priority theme and publication period. **A various research themes in hypospadiology based on the recent global international Delphi survey, in order to gain a clearer understanding of the distribution of efforts and literature concerning these highly prioritized research themes covering etiology, tissue engineering, pre-clinical models, device/technology evaluation, phenotyping, surgical techniques, surgical training and postoperative outcomes. (Figure 3A)**

**Figure 3.**
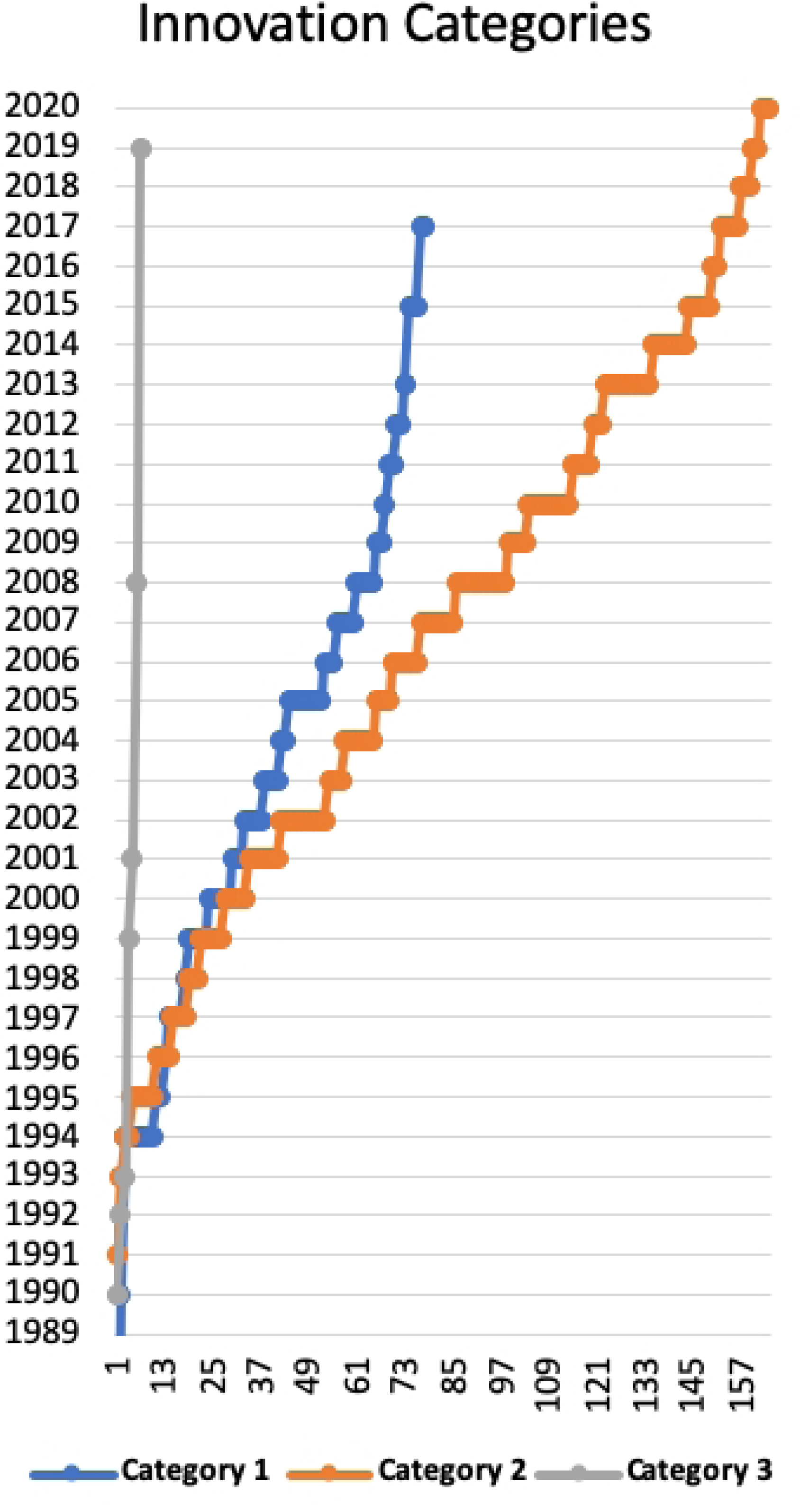
Subgroup analyses of hypospadias research (1990–2023). (A) Distribution of studies by research themes. (B) Keyword frequency analysis indicating major areas of research emphasis and identifying potential gaps in the hypospadias and urologic surgery literature. (C) Distribution of journal publications contributing to the field over the study period.

The analysis revealed that the majority of studies focused on surgical techniques (30.5%), etiology (25.8%), and postoperative outcomes (22.6%), reflecting a strong emphasis on clinical and procedural aspects of hypospadias care. Phenotyping represented 10.5% of the literature, indicating growing interest in personalized approaches. In contrast, surgical training (2.6%), tissue engineering (2.1%), pre-clinical models (0.5%), and device/technology evaluation (0%) were significantly underrepresented, highlighting important gaps in innovation, education, and translational research.

### 3.4 Keywords Co-Occurrences and journals

The frequency of keywords highlights key areas of focus and research gaps in hypospadias and urologic surgery. Dominant topics like hypospadias (197), surgery (94), and urethra (57) emphasize the importance of surgical techniques and urethral reconstruction. High frequencies of postoperative complications (52) and reoperation (28) reflect ongoing challenges in achieving optimal outcomes. Lower frequencies of genetics (17), urethral diseases (13), and endocrine disruptors (9) point to underexplored areas that could shed light on hypospadias etiology. Niche topics like suture techniques (12), plastic surgery (5), and urethroplasty (4) suggest opportunities to refine surgical practices, while emerging areas like skin transplantation (7), laser therapy (4), and penile erection (4) could offer innovative treatment options. Addressing these gaps would advance scientific understanding and improve patient outcomes. **(Figure 3B)**

The distribution of journal publications reveals a strong concentration of articles in prominent urology-related journals, with Journal of Urology leading with 74 publications, followed by Journal of Pediatric Urology with 19. This highlights the focus on pediatric urology, specifically, as a key area of research. Other urology-focused journals, such as BJU International (12) and Urology (11), also contribute significantly to the field. These numbers suggest that the research on topics like hypospadias and urologic procedures is predominantly published in specialized urology and pediatric surgery journals. **(Figure 3C)**

### 3.5 Quality Assessment and Risk of Bias

The quality assessment of the 20 included studies, based on their study design and the appropriate risk of bias tools, is summarized in **Supplementary Table 1**. Overall, most studies were rated as good in quality, reflecting well-defined methodologies and comprehensive reporting. However, one study was rated as fair due to insufficient information regarding selection bias, including unclear randomization methods and incomplete outcome data in certain sections. In contrast, studies rated as poor were typically observational studies with more than 20% loss to follow-up and critical methodological limitations, such as inadequate patient selection criteria and incomplete reporting of adverse events.

### 3.6 Temporal and Thematic Evolution of Surgical Innovation

To further explore thematic trends and the nature of innovation within hypospadias research, the top 200 most-cited articles wre categorized into three innovation categories: Category 1 (Surgical Technique Innovation), Category 2 (Etiological aspects, Perioperative Management, Outcome Assessment & Long-term Follow-up), and Category 3 (Technology, Materials & Tissue Engineering). Temporal analysis revealed that Category 1 articles appeared earliest, with most originating in the 1990s and early 2000s. These focused on procedural modifications, such as new flap techniques or urethral plate preservation strategies. In contrast, Category 2 articles became more prominent in the 2010s, particularly among those with high Disruption Index values (DI ≥ 0.5), indicating their role in shifting research focus toward patient-centered outcomes, complication tracking, and long-term follow-up. Category 3 was the least represented, reflecting a current gap in innovation related to technology integration, material science, and translational applications such as tissue engineering. This thematic evolution is visualized in **Figure 4**, which maps the distribution of innovation categories across decades and highlights the relative DI scores of each article.

**Figure 4:**
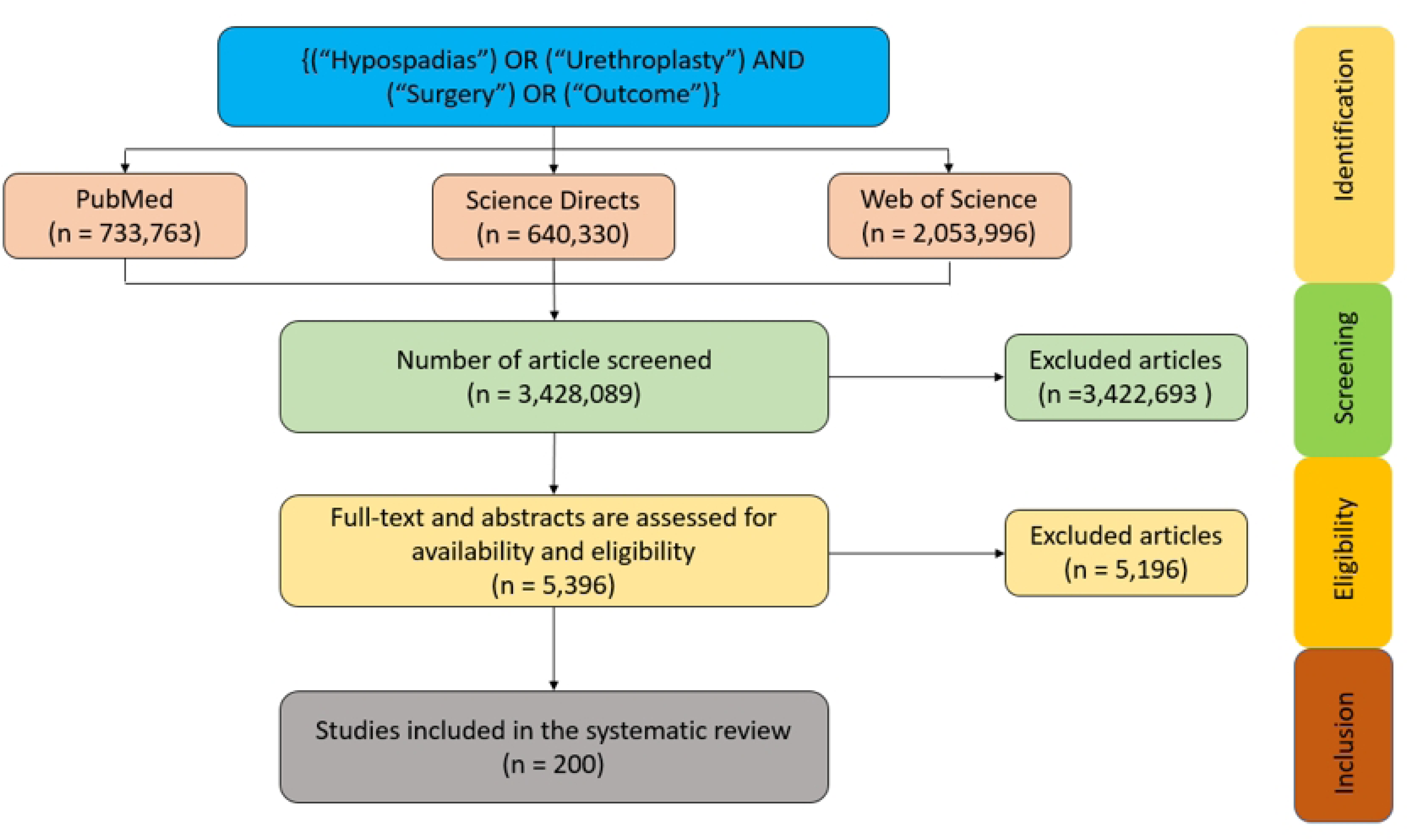
Temporal analysis of innovation categories over the studies with highest disruption index. (X-axis represent the number of the studies in the specified category).

## 4. DISCUSSION

To our knowledge, this is the first study to apply the Disruption Index (DI) to hypospadiology, identifying disruptive publications and evolving trends in the field. The analysis offers a comprehensive overview of hypospadias and urethroplasty research from 2000 to 2023, revealing significant growth in both publication volume and impact. This section discusses the key trends, strengths, limitations, and implications for future research in hypospadias and urethroplasty.

In a biobiographic analysis on 2021, [Dogan & Ipek]The top 10 authors who made the highest contribution to the literature about hypospadias were Nordenskjold A, Hayashi Y, Baskin LS, Kohri K, Kojima Y, Mizuno K, Duckett JW, Nonomura K, Snodgrass W, and Carmichael SL. While measuring the disruption index. Several bibliometric studies have analyzed highly cited publications on hypospadias to identify key research trends and influential works in the field. [10-12] Our study enhances previous work by integrating the Disruption Index (DI), revealing how publications shape future research. Studies with high DI values typically introduced novel surgical techniques, innovative training, or comprehensive reviews, making a lasting impact on both academic and clinical practices. While traditional citation metrics reflect academic attention, they don’t distinguish between incremental and transformative work. The DI, however, assesses how much a publication shifts the direction of future research, highlighting paradigm-changing contributions.

In hypospadias research, the Disruption Index (DI) highlights studies that significantly redirect academic discourse or clinical practice. While the DI offers a deeper understanding of influence beyond citation volume, it has limitations, including reduced interpretability for recent publications and sensitivity to database coverage. Citation count, though a standard measure of academic impact, doesn’t differentiate between reinforcing existing paradigms and truly disruptive work. Citation biases, such as self-citations and the age of publications, can also skew results. This study demonstrates the utility of the DI in hypospadias research, providing insights into shifting paradigms despite its limitations. The Disruption Index (DI) addresses citation limitations by measuring how a publication influences future research. It distinguishes between studies that build on a focal paper independently (indicating disruption) and those that rely on prior work (indicating consolidation). The DI highlights transformative studies that shift scientific inquiry, offering a richer perspective on research impact when used alongside citation counts. This helps identify seminal works that may be underrecognized by citation volume alone but have profoundly shaped the field.

Thematic analysis revealed that nearly 80% of studies focused on surgical techniques, etiology, and postoperative outcomes, reflecting the field’s traditional emphasis on refining operative strategies and understanding hypospadias causes and outcomes. However, there is limited attention to emerging areas like phenotyping, surgical training, tissue engineering, and pre-clinical models, highlighting a critical gap. Notably, no highly cited studies addressed device or technology evaluation, despite the growing relevance of digital tools in surgical planning and follow-up. This suggests that hypospadias research has not fully aligned with broader trends in pediatric surgery and biomedical innovation, such as simulation, regenerative medicine, and personalized approaches. Addressing these gaps could help shape a more future-oriented research agenda, improving surgical quality and patient outcomes. A significant portion of studies were categorized under “Others,” indicating interdisciplinary research in niche journals. Journals outside of urology, such as the Journal of Pediatrics and the British Journal of Anaesthesia, have contributed valuable perspectives. While urology and pediatric urology journals remain key, enhancing interdisciplinary collaboration across fields like pediatrics, plastic surgery, and environmental health could expand the reach and impact of hypospadias research. Focusing on underrepresented journals could further influence areas that intersect with pediatric urology and surgical care.

This study provides valuable insights into hypospadias and urethroplasty research, but several limitations should be noted. First, our analysis was limited to English-language publications, potentially excluding relevant studies in other languages. Second, citation-based metrics (e.g., DI, total citations) may not fully capture the clinical relevance or quality of a study, as citation counts can be influenced by factors like journal prestige or author reputation. Additionally, the quality of included studies varied, with some observational studies and systematic reviews showing moderate or high bias, which may have affected our conclusions.

The inclusion of the Disruption Index (DI) adds a key dimension to understanding the field’s evolution. While traditional citation metrics highlight popular studies, the DI identifies works that have reshaped research trajectories. Interestingly, some studies with low citation counts had high disruption scores, indicating their impact in redirecting research focus rather than simply accumulating citations. This highlights the importance of considering both citation volume and impact type in assessing progress in evolving fields like hypospadiology.

## 5. Conclusion

This study offers a comprehensive bibliometric and disruption-based analysis of hypospadias surgery, highlighting key trends, influential publications, and emerging topics. The Disruption Index provides unique insights into how studies have reshaped the field, complementing traditional citation metrics. Our findings show a strong focus on surgical techniques, etiology, and outcomes, with gaps in areas like surgical training, tissue engineering, and device innovation. Addressing these gaps is crucial for fostering innovation, improving education, and integrating new technologies into clinical practice. Combining disruption metrics with traditional bibliometrics can guide future research and accelerate advances in hypospadias care and urologic surgery.

## Artificial Intelligence and Machine Learning Declaration

This study did not involve the use of artificial intelligence (AI) or machine learning (ML) techniques in the data collection, analysis, or interpretation phases. All bibliometric analyses and calculations, including the Disruption Index, were performed using established bibliometric databases and standard statistical methods.”

## Ethical Approval

Not applicable as this is a review study

## Data Availability

The minimal data set is available upon request

## Acknowledgement

The authors would like to express their gratitude to all individuals and institutions for their valuable collaboration, insight, and support.

## Table Legends

- Table 1. Details of publications with the top 20 DI scores
- Supplemental Table 1. Quality Assessment of Included Studies Based on Study Design and Risk of Bias Tools

